# Associations between gestational weight gain adequacy and neonatal outcomes in Tanzania

**DOI:** 10.1101/2021.08.19.21262273

**Authors:** Nandita Perumal, Dongqing Wang, Anne Marie Darling, Molin Wang, Enju Liu, Willy Urassa, Andrea Pembe, Wafaie W. Fawzi

## Abstract

**Introduction:** Gestational weight gain (GWG) is associated with fetal and newborn health; however, data from sub-Saharan Africa are limited.

**Methods:** We used data from a prenatal micronutrient supplementation trial among a cohort of HIV-negative pregnant women in Dar es Salaam, Tanzania to estimate the relationships between GWG and newborn outcomes. GWG adequacy was defined as the ratio of the total observed weight gain over the recommended weight gain based on the Institute of Medicine body mass index (BMI)-specific guidelines. Newborn outcomes assessed were: stillbirth, perinatal death, preterm birth, low birthweight, macrosomia, small-for-gestational age (SGA), large-for-gestational age (LGA), stunting at birth, and microcephaly. Modified Poisson regressions with robust standard error were used to estimate the relative risk of newborn outcomes as a function of GWG adequacy.

**Results:** Of 7561 women included in this study, 51% had severely inadequate (<70%) or inadequate GWG (70-90%), 31% had adequate GWG (90-125%), and 18% had excessive GWG (≥125%). Compared to adequate GWG, severely inadequate GWG was associated with a higher risk of low birthweight, SGA, stunting at birth, and microcephaly; whereas excessive GWG was associated with a higher risk of LGA and macrosomia.

**Conclusion:** Interventions to support optimal gestational weight gain are needed and are likely to improve newborn outcomes.

## INTRODUCTION

Weight gain during pregnancy is an important predictor of maternal pregnancy outcomes and newborn health.[1] The 2009 Institute of Medicine (IOM) guidelines provide specific recommendations for total gestational weight gain (GWG) during pregnancy according to a woman’s pre-pregnancy body-mass-index (BMI).[1] Inadequate or excessive GWG has been associated with a wide range of adverse maternal and neonatal health outcomes.[1,2] For example, inadequate GWG has been associated with a higher risk of low birthweight, small-for-gestational age (SGA), and preterm birth, whereas excessive GWG has been associated with a higher risk of cesarean delivery, large-for-gestational age (LGA), and macrosomia.[1–3] Adverse birth outcomes in turn are associated with an increased risk of mortality,[4] suboptimal growth,[5] lower neurodevelopment[6] and schooling achievement,[7] and adverse cardiometabolic outcomes later in life. Gaining weight during pregnancy within the recommended guidelines is therefore important to minimize the risk of adverse birth outcomes.

The IOM recommendations for GWG, however, are based on data from high-income settings and do not include evidence from any studies examining the relationship between GWG and neonatal outcomes in low-income settings. In high-income countries, the prevalence of adverse birth outcomes is low (∼4-5%) and maternal overweight and excessive GWG are a larger concern during antenatal care.[3,8–10] In contrast, 25% of all live births globally that are estimated to be low birthweight or preterm occur in countries in sub-Saharan Africa.[8,9] In addition, evidence from population-representative surveys in 67 low and middle-income countries (LMICs) suggests that women in sub-Saharan Africa on average have an estimated total GWG of 6.6 kg (95% uncertainty intervals: 3.4, 9.9), which is less than 60% of the minimum recommended GWG for normal weight women based on the IOM guidelines.[11]

A recent systematic review and meta-analysis of a relatively few studies conducted in sub-Saharan Africa suggests that inadequate and excessive GWG are similarly associated with low birthweight and macrosomia, respectively, as observed in high-income settings. However, most of the studies included in the meta-analysis were cross-sectional or retrospective cohort study design, were based on a sample size of <500 participants, were noted to have poor control of confounding variables, and examined relationships primarily with newborn weight.[12] In this study, we use data from a large, prospective cohort of pregnant women in Dar es Salaam, Tanzania, to examine the relationships between GWG during pregnancy and a wide range of neonatal outcomes. In addition, we assessed whether associations between maternal GWG and neonatal outcomes were modified by maternal age and nutritional status, as assessed by maternal pre-pregnancy BMI.

## METHODS

### Study design and population

We used secondary data from participants enrolled in a double-blind randomized controlled trial of daily prenatal multiple micronutrient supplementation during pregnancy conducted between 2001 to 2005 in Dar es Salaam, Tanzania. The trial procedures and primary findings are published in detail elsewhere.[13] Briefly, 8428 pregnant women between 12 to 28 weeks gestation were randomized to receive either a daily multiple micronutrient supplement (MMS) or placebo during pregnancy to investigate the effects on perinatal outcomes, including low birthweight (<2500 grams), preterm birth, and fetal death. In line with standard of care in Tanzania, all participants received daily supplements of 0.25 mg folic acid, 60 mg of ferrous sulfate, and sulfadoxine-pyrimethamine for malaria prophylaxis.

Participants were screened and recruited from antenatal care clinics in nine health centers in Dar es Salaam, Tanzania, if they met the following eligibility criteria: (i) tested negative for human immunodeficiency virus (HIV) infection, (ii) were between 12 and 27 weeks of gestational age based on date of last menstrual period, (iii) were 18 years of older, and (iv) intended to stay in Dar es Salaam for at least 1 year after delivery. Participants were followed monthly from the time of enrolment until 32 weeks gestational age, after which they were followed every 2 weeks until the 36^th^ week of gestation, and then every week until 6 weeks postpartum. Trained research nurses administered the questionnaires to collect socio-demographic information, detailed medical and obstetric history, and performed clinical examinations at baseline and each follow-up visit. Written informed consent was obtained from all participants enrolled in the trial.

### Maternal weight and gestational weight gain assessment

Maternal weight was measured at enrollment and at every scheduled follow-up visit. Weight was recorded to the nearest 100 grams with balance scales with women wearing light clothing without shoes. To calculate the observed GWG, we subtracted the women’s first trimester weight from the last observed weight prior to delivery.

Because women were eligible to be in the trial if they were between 12 to 27 weeks of age, we did not have first trimester weight for 98% of the participants. We therefore imputed the first trimester weight for all women at 9 weeks gestation (mid-point of the first trimester), as a proxy of pre-pregnancy weight, using a mixed-effects model with restricted cubic splines with 3 knots using all available maternal weights during the first and second trimesters. The methodology and validation procedures for the imputation have been described in detail elsewhere.[14] We then used the imputed or observed first trimester weight for each participant to as a proxy to assess maternal pre-pregnancy BMI and classified women as being underweight (BMI <18.5 kg/m^2^), normal weight (BMI 18.5 to 24.9 kg/m^2^), overweight (BMI 25 to 29.9 kg/m^2^) or obese (BMI ≥30 kg/m^2^) based on the World Health Organization criteria. For participants who were <20 years of age at enrolment, we used the World Health Organization BMI-for-age growth references [15] to classify adolescents as underweight (<-2 SD), normal weight (−2 SD to <1 SD), overweight (1 SD to <+2 SD) or obese (≥+2 SD).

We then used GWG adequacy ratio based on IOM recommendations as the primary exposure of interest. This metric has been previously used in studies of GWG[16] and offers the major advantage of being independent of gestational duration as it is the ratio of absolute maternal weight gain over the recommended weight gain in the same gestational duration, estimated as follows:

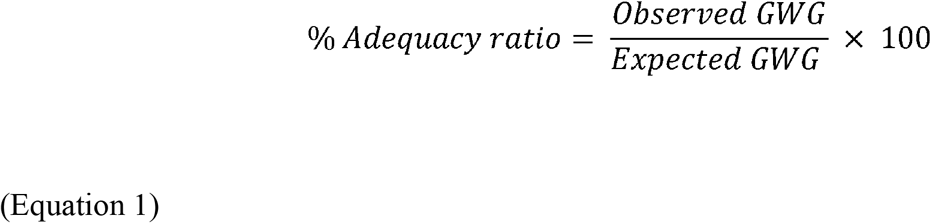

Where, the recommended weight gain according to IOM guidelines is estimated as follows:

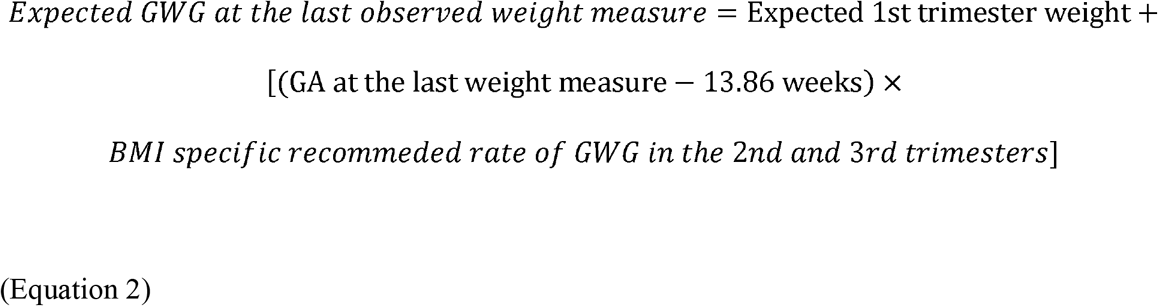

Where the expected weight gain for each participant was the sum of the BMI-specific expected 1st trimester weight (at the end of 13^6/7^ weeks) and the weight gain up to last weight measure based on the BMI-specific expected rate of weight gain in the 2nd and 3rd trimesters (Table 1). The GWG adequacy ratio was derived as a continuous measure and categorized as follows for analysis: severely inadequate (<70%); inadequate (70 to <90%), adequate (90 to <125%), and excessive (≥125%).

**Table 1:** Institute of Medicine guidelines for expected weight gain and rate of weight gain based on maternal pre-pregnancy body mass index (BMI) classification.

| **BMI classifications** | **Expected 1st trimester weight (13<sup>6/7</sup> weeks)** | **Recommended rate of weight gain in 2<sup>nd</sup> and 3<sup>rd</sup> trimester** |
| --- | --- | --- |
| Underweight (<18.5 kg/m<sup>2</sup>) | 2 kg | 0.51 kg/week |
| Normal weight (18.5 to 24.9 kg/m<sup>2</sup>) | 1 kg | 0.42 kg/week |
| Overweight (25 to 29.9 kg/m<sup>2</sup>) | 0.5 kg | 0.28 kg/week |
| Obese (≥30 kg/m<sup>2</sup>) | 0.5 kg | 0.22 kg/week |

We also used the INTERGROWTH-21^st^ Gestational Weight Gain standards (IG-GWG) to derive standardized indices (z-scores) of GWG for normal weight women.[17] The IG-GWG provide a new reference population for assessing GWG adequacy associated with optimal maternal and perinatal outcomes as these standards are based on a multi-ethnic population of healthy normal weight pregnant women from high-socioeconomic status and with no known risk factors. We derived GWG z-scores for each participant and outlying values, defined as observations with GWG z-score above 6 standard deviations (SD) or below -6 SD, were excluded from analysis. GWG z-scores were categorized into four categories for analysis (<-2 SD, -2 SD to <-1 SD, -1 SD to <1 SD, and ≥1 SD).

### Outcome assessment

We assessed several neonatal mortality and anthropometric outcomes. Detailed information on infant status at birth and up to 6 weeks postpartum were collected during scheduled visits. Neonatal mortality outcomes examined were stillbirth, defined as death of the fetus after 28 weeks; and perinatal death, defined as death of an infant after 28 weeks gestation and within the first 7 days of life. Trained research midwives measured newborn weight at the time of delivery to the nearest 10 grams using digital scales and newborn length using a length board. We used standard definitions of preterm birth (<37 weeks gestational age), low birthweight (<2500 grams), and macrosomia (>4000 grams). We used the INTERGROWTH-21^st^ Newborn Size Standards to derive gestational age- and sex-standardized percentiles/z-scores to classify: small-for-gestational age (SGA; <10^th^ percentile of birthweight), large-for-gestational age (LGA; >90^th^ percentile for birthweight), stunting at birth (length-for-gestational age z-score <-2SD), and microcephaly (head circumference-for-gestational age z-score <-2SD).

### Confounders

We identified several confounders of the associations between GWG adequacy and neonatal outcomes. These included maternal age (<20, 20-24, 25-29, or ≥30 years), education (0-4, 5-7, 8-11, or ≥12 years), marital status (living alone or with partner), parity (none, 1, 2, or ≥3), wealth index, smoking status (yes/no), maternal alcohol consumption (never, <1 per week, or ≥1 per week), malaria infection at enrolment (yes/no), and prenatal supplementation group (multiple micronutrients vs placebo). Wealth index was constructed by a linear index of asset ownership indicators (television, refrigerator, radio, sofa, and fan) based on principal component analysis.[18] We did not adjust for maternal reproductive history as adjusting for these factors has been shown to introduce bias and underestimate the relationships for factors in current pregnancy.[19] We adjusted for maternal pre-pregnancy BMI as a confounder as well, but did not adjust for any factors, such as gestational diabetes or hypertension, that would fall on the causal pathway from GWG to neonatal outcomes (Figure 1).

**Figure 1:**
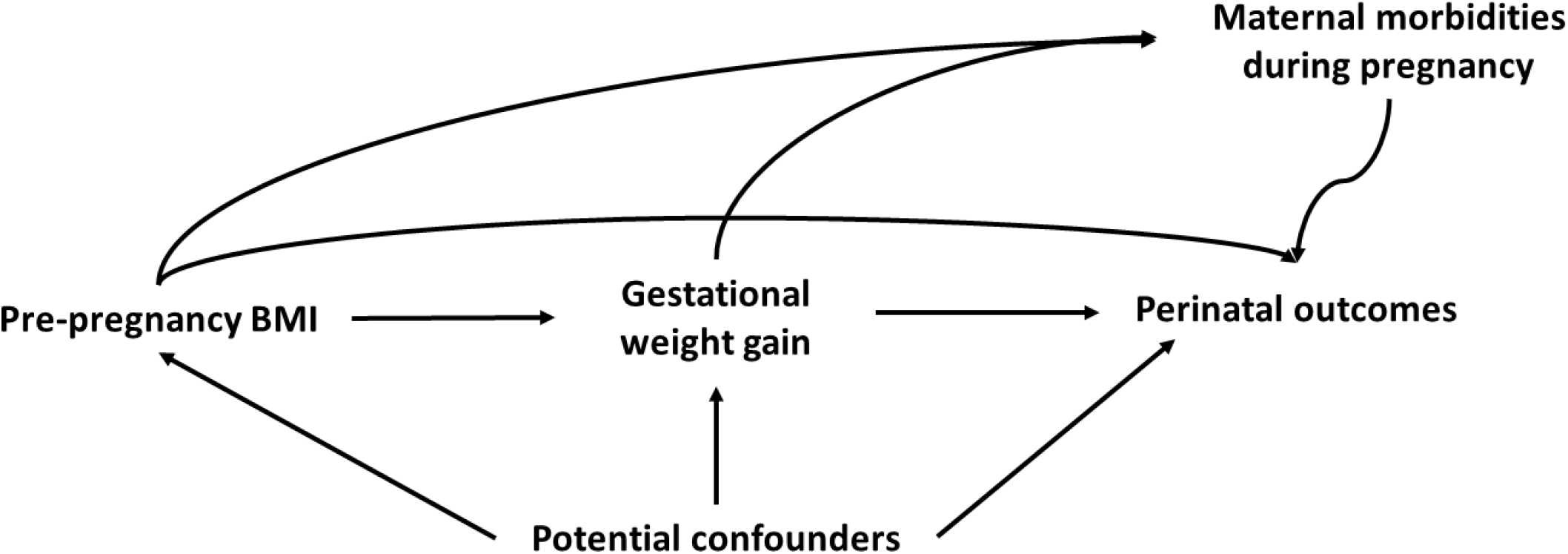
Conceptual framework for the association between maternal gestational weight gain and perinatal outcomes.

### Statistical analysis

Analyses were limited to women with singleton pregnancy (n = 156 women with twins or triplets excluded) and known date of gestational age by last menstrual period. We further excluded weight measures (but not participants) if the weight was taken after 43 weeks gestational age as these were likely to be postpartum weight (n = 140 weight measures), and if the values were extreme (defined as <30 kg or >120 kg; n = 7 weight measures). We used modified Poisson regression with robust standard error to estimate the multivariable adjusted relative risks of neonatal outcomes as a function of GWG adequacy in primary analysis, and GWG z-scores in secondary analysis. Because a participant’s last weight measure may be in the second or third trimester, we base inferences regarding total GWG adequacy on participants for whom at least one weight measure in the third trimester was available. We further assessed relationships with ‘early’ GWG adequacy using the last weight measure in the second trimester. We used two-way interaction terms with GWG adequacy to assess whether relationships between GWG adequacy and neonatal outcomes were modified by maternal pre-pregnancy BMI and maternal age. Due to the small sample size of women classified as having obesity based on their pre-pregnancy BMI (n = 379), we combined women with overweight and obese pre-pregnancy BMI classification for interaction analyses. Wald test for interaction was considered statistically significant at two-sided alpha <0.05. All analyses were conducted in Stata 16 (College Station, Texas).

## RESULTS

Data from 7561 mother-infant dyad were included in this study. The mean (SD) gestational age of participants at enrolment was 21 (3.4) weeks and at delivery was 39 (3.6), with an average of 4.2 weight measures taken per participant during pregnancy. Most participants were between 20-30 years of age, had less than or equal to primary school education, were married, were primiparous, and were normal weight (Table 2). Overall, 25% of women had either severely inadequate GWG, 26% had inadequate GWG, 31% had adequate GWG, and 18% gained excessive GWG. The prevalence of stillbirths (3.5%), perinatal death (6.1%), LBW (6.8%), macrosomia (2.5%), and microcephaly (8.5%) were relatively low; however, SGA (20%), LGA (13%), stunting at birth (26%), and preterm birth (17%) prevalence were high.

**Table 2:** Participant characteristics for women in the Prenatal Vitamins Trial.

| Characteristic | Gestational weight gain adequacy |  |  |  |  | P<sup>1</sup> |
| --- | --- | --- | --- | --- | --- | --- |
|  | Overall<br>(n = 7561) | Severely<br>inadequate<br>(<70%)<br>(n = 1928) | Inadequate<br>(70 to <90%)<br>(n = 1938) | Adequate<br>(90 to <125%)<br>(n = 2315) | Excessive<br>(≥ 125%)<br>(n = 1392) |  |
| **Maternal age<sup>2</sup>, yrs** |  |  |  |  |  |  |
| <20 | 1216 (16) | 368 (19) | 313 (16) | 371 (16) | 164 (12) | <0.001 |
| 20-24 | 3002 (40) | 793 (42) | 823 (43) | 890 (39) | 496 (36) |  |
| 25-29 | 2027 (27) | 441 (23) | 489 (25) | 652 (28) | 445 (32) |  |
| ≥30 | 1279 (17) | 305 (16) | 300 (16) | 388 (17) | 286 (21) |  |
| **Education<sup>3</sup>, yrs** |  |  |  |  |  |  |
| 0 to 4 yrs, | 841 (11) | 251 (13) | 253 (13) | 221 (9.60) | 116 (8.34) | <0.001 |
| 5 to 7 yrs | 5009 (67) | 1344 (70) | 1283 (67) | 1519 (66) | 863 (62) |  |
| 8 to 11 yrs | 1288 (17) | 249 (13) | 315 (16) | 428 (19) | 296 (21) |  |
| 12+ yrs | 388 (5.2) | 64 (3.35) | 75 (3.89) | 133 (5.78) | 116 (8.34) |  |
| **Married or living with partner<sup>4</sup>** | 6632 (88) | 1674 (88) | 1700 (89) | 2025 (88) | 1233 (89) | 0.97 |
| **Prior pregnancies<sup>5</sup>** |  |  |  |  |  |  |
| None | 3452 (46) | 901 (47) | 891 (46) | 1066 (46) | 588 (42) | 0.002 |
| 1 | 2088 (28) | 507 (27) | 535 (28) | 662 (29) | 380 (27) |  |
| 2 | 1099 (15) | 254 (13) | 271 (14) | 316 (14) | 256 (18) |  |
| ≥ 3 | 893 (12) | 241 (13) | 229 (12) | 257 (11) | 166 (12) |  |
| **Supplementation group** |  |  |  |  |  |  |
| Placebo | 3772 (50) | 1005 (53) | 965 (50) | 1131 (49) | 671 (48) | 0.04 |
| MMS | 3789 (50) | 909 (47) | 969 (50) | 1184 (51) | 727 (52) |  |
| **Wealth quartile, n (%)<sup>6</sup>** |  |  |  |  |  |  |
| 1<sup>st</sup> (lowest) | 2964 (39) | 894 (47) | 792 (41) | 834 (36) | 444 (32) | <0.001 |
| 2<sup>nd</sup> | 1045 (14) | 293 (15) | 282 (15) | 306 (13) | 164 (12) |  |
| 3<sup>rd</sup> | 2109 (28) | 478 (25) | 539 (28) | 682 (30) | 410 (29) |  |
| 4<sup>th</sup> (highest) | 1420 (19) | 244 (13) | 315 (16) | 487 (21) | 374 (27) |  |
| **Maternal pre-pregnancy BMI, n (%)** |  |  |  |  |  |  |
| Underweight (<18.5 kg/m<sup>2</sup>) | 797 (11) | 285 (15) | 292 (15) | 182 (7.86) | 38 (2.72) | <0.001 |
| Normal weight (18.5 – 24.9 kg/m<sup>2</sup>) | 4962 (66) | 1390 (73) | 1477 (76) | 1698 (73) | 397 (28) |  |
| Overweight (25 – 29.9 kg/m<sup>2</sup>) | 1423 (19) | 193 (10) | 141 (7.29) | 383 (17) | 706 (51) |  |
| Obese (≥ 30 kg/m<sup>2</sup>) | 379 (5.01) | 46 (2.40) | 24 (1.24) | 52 (2.25) | 257 (18) |  |
| **Maternal height** |  |  |  |  |  |  |
| Height <155 cm | 3439 (45) | 954 (50) | 910 (47) | 990 (43) | 585 (42) | <0.001 |
| Height ≥155 cm | 4122 (55) | 960 (50) | 1024 (53) | 1325 (57) | 813 (58) |  |
| **Maternal alcohol consumption, n (%)<sup>7</sup>** |  |  |  |  |  |  |
| Never | 6584 (87) | 1688 (88) | 1712 (89) | 2011 (87) | 1173 (84) | 0.002 |
| Less than once per week | 712 (9.45) | 173 (9.05) | 150 (7.80) | 221 (9.57) | 168 (12) |  |
| Once or more times per week | 238 (3.16) | 50 (2.62) | 62 (3.22) | 78 (3.38) | 48 (3.46) |  |
| **Maternal malaria infection<sup>8</sup>, n (%)** | 111 (1.5) | 35 (1.92) | 27 (1.46) | 31 (1.40) | 18 (1.35) | 0.49 |
| **Maternal reproductive history<sup>9</sup>** |  |  |  |  |  |  |
| Previous abortions or miscarriages, n (%) | 1108 (16) | 256 (15) | 265 (15) | 340 (16) | 247 (19) | 0.008 |
| Previous stillbirth, n (%) | 221 (3.30) | 55 (3.30) | 53 (3.08) | 61 (2.96) | 52 (4.17) | 0.27 |
| Previous fetal loss, n (%) | 1296 (17) | 303 (16) | 310 (16) | 394 (17) | 289 (21) | 0.001 |

<sup>1</sup> P-value for Chi-2 statistic for difference between categories; <sup>2</sup> Maternal age was missing for n=37 participants; <sup>3</sup> Maternal education was missing for n=35 participants; <sup>4</sup> Marital status missing for 35 participants; <sup>5</sup> Maternal parity was missing for 41 participants; <sup>6</sup> Wealth quartile was missing for 23 participants; <sup>7</sup> Maternal alcohol consumption was missing for 39 participants; <sup>8</sup> Maternal malaria infection at baseline was missing for 301 participants; <sup>9</sup> Maternal reproductive history was missing for 757 and 874 participants for history of abortions and stillbirths, respectively.

The relationships between GWG adequacy and neonatal outcomes are summarized in Table 3. Compared to adequate GWG, total severely inadequate GWG was associated with a higher risk of LBW (adjusted RR 1.64, 95% confidence intervals (CI): 1.24, 2.16), SGA (RR 1.81, 95%CI: 1.53, 2.14), stunting at birth (RR 1.27, 95%CI: 1.06, 1.52), and microcephaly (RR 1.35, 95%CI: 1.04, 1.74); but a lower risk of LGA (RR 0.69, 95%CI: 0.56, 0.86) and macrosomia (RR 0.29, 95%CI: 0.16, 0.54). Total inadequate GWG was also associated with a higher risk of SGA and microcephaly, and a lower risk of LGA (Table 3). Total excessive GWG was positively associated with the risk of stillbirth (RR 1.60, 95%CI: 1.03, 2.42) and LGA (RR 1.44, 95%CI: 1.14, 1.81). GWG adequacy in the second trimester was similarly associated with neonatal outcomes, although confidence intervals crossed the null for some associations. Notably, severely inadequate GWG in the second trimester was positively associated with the risk of preterm birth (RR 1.58, 95%CI: 1.31, 1.91) and excessive GWG in the second trimester was positively associated with the risk of macrosomia (RR 1.98, 95%CI: 1.27, 3.08) (Table 3).

**Table 3:** Associations between percent adequacy of gestational weight gain (GWG) compared to the Institute of Medicine 2009 recommendations and neonatal outcomes^1^.

| Characteristic | Total<br>n | No. of<br>cases (%) | Severely<br>Inadequate (<70%)<br>RR (95% CI) | Inadequate<br>(70% to <90%)<br>RR (95% CI) | Adequate<br>(90% to 125%)<br>Reference | Excessive<br>(>125%)<br>RR (95% CI) |
| --- | --- | --- | --- | --- | --- | --- |
| **Total GWG** |  |  |  |  |  |  |
| Stillbirth | 6578 | 209 (3.18) | 1.16 (0.79, 1.71) | 0.97 (0.64, 1.46) | 1.0 | 1.60 (1.03, 2.42) |
| Perinatal death | 6375 | 350 (5.49) | 1.04 (0.77, 1.40) | 0.85 (0.62, 1.17) | 1.0 | 1.23 (0.89, 1.69) |
| Preterm-born | 6370 | 901 (14.2) | 1.12 (0.93, 1.36) | 1.00 (0.83, 1.22) | 1.0 | 1.23 (0.97, 1.56) |
| Low birthweight | 6205 | 381 (6.14) | 1.64 (1.24, 2.16) | 1.17 (0.87, 1.57) | 1.0 | 1.30 (0.89, 1.89) |
| Small-for-gestational age | 6188 | 1248 (20) | 1.81 (1.53, 2.14) | 1.47 (1.24, 1.75) | 1.0 | 0.85 (0.67, 1.07) |
| Large-for-gestational age | 6188 | 765 (12.4) | 0.69 (0.56, 0.86) | 0.76 (0.61, 0.93) | 1.0 | 1.44 (1.14, 1.81) |
| Stunting at birth | 4590 | 1226 (27) | 1.27 (1.06, 1.52) | 1.20 (1.00, 1.43) | 1.0 | 0.97 (0.78, 1.20) |
| Microcephaly | 5435 | 472 (8.69) | 1.35 (1.04, 1.74) | 1.32 (1.02, 1.71) | 1.0 | 0.91 (0.65, 1.26) |
| Macrosomia | 6205 | 160 (2.58) | 0.29 (0.16, 0.54) | 0.65 (0.41, 1.03) | 1.0 | 1.52 (1.00, 2.31) |
| **Early GWG (2<sup>nd</sup> trimester)** |  |  |  |  |  |  |
| Stillbirth | 7054 | 247 (3.50) | 1.05 (0.70, 1.57) | 1.02 (0.72, 1.46) | 1.0 | 1.24 (0.83, 1.86) |
| Perinatal death | 6750 | 407 (6.03) | 1.04 (0.76, 1.42) | 0.89 (0.67, 1.19) | 1.0 | 1.06 (0.77, 1.46) |
| Preterm-born | 6807 | 1075 (16) | 1.58 (1.31, 1.91) | 0.89 (0.75, 1.07) | 1.0 | 0.99 (0.79, 1.24) |
| Low birthweight | 6579 | 448 (6.81) | 1.67 (1.27, 2.21) | 1.17 (0.90, 1.51) | 1.0 | 1.09 (0.77, 1.55) |
| Small-for-gestational age | 6559 | 1310 (20) | 1.27 (1.06, 1.53) | 1.36 (1.16, 1.60) | 1.0 | 0.86 (0.70, 1.07) |
| Large-for-gestational age | 6559 | 874 (13.3) | 1.22 (0.99, 1.51) | 0.78 (0.63, 0.95) | 1.0 | 1.19 (0.95, 1.50) |
| Stunting at birth | 4741 | 1256 (26) | 0.99 (0.81, 1.21) | 1.05 (0.89, 1.25) | 1.0 | 1.08 (0.88, 1.33) |
| Microcephaly | 5615 | 479 (8.53) | 1.02 (0.76, 1.35) | 1.01 (0.79, 1.29) | 1.0 | 1.02 (0.75, 1.38) |
| Macrosomia | 6579 | 166 (2.52) | 0.42 (0.21, 0.81) | 0.63 (0.38, 1.02) | 1.0 | 1.98 (1.27, 3.08) |

<sup>1</sup> Multivariable models adjusted for maternal age, education, marital status, wealth quintile, smoking status, alcohol consumption, maternal pre-pregnancy body-mass-index, parity, malaria infection, and supplementation group assignment.

The relationships between GWG adequacy and neonatal outcomes differed by maternal pre-pregnancy BMI (Figure 2; Supplemental Tables 1-4). For example, severely inadequate GWG was associated with a substantially higher risk of LBW among underweight women (RR 2.95, 95%CI: 1.95, 4.48) compared to normal weight women (RR 1.34, 95%CI: 0.97, 1.87; P-for-interaction = 0.03) (Supplementary Table 1). Similarly, excessive GWG was associated with a higher risk of stillbirth among underweight women (RR 3.13, 95%CI: 0.96, 10.2) compared to normal weight women (RR 1.73, 95%CI: 0.95, 3.15; P-for-interaction = 0.04) (Supplementary Table 1). Among overweight or obese women, severely inadequate and excessive GWG were associated with a higher risk of perinatal death (RR 1.87, 95%CI: 1.06, 3.30 and RR 1.84, 95%CI: 1.09, 3.12, respectively); however, these relationships were not statistically significantly different from relationships between GWG adequacy and perinatal death among normal weight women (Supplementary Table 1). We also observed some differences in the relationships between GWG adequacy by maternal age (Supplementary Table 3). Severely inadequate GWG was more strongly associated with the higher risk of stunting at birth (RR 1.39, 95%CI: 1.03, 1.89) and with a lower risk macrosomia (RR 0.45, 95%CI: 0.13, 1.54) among women who were >30 years, compared with women 20-29 years of age (stunting: RR 0.95, 95%CI: 0.72, 1.26; P-for-interaction =0.01; macrosomia: RR 0.69, 95%CI: 0.27, 1.77; P-for-interaction = 0.03). Other relationships between GWG adequacy and neonatal outcomes were not modified by maternal age (Supplementary Tables 3 and 4).

**Figure 2:**
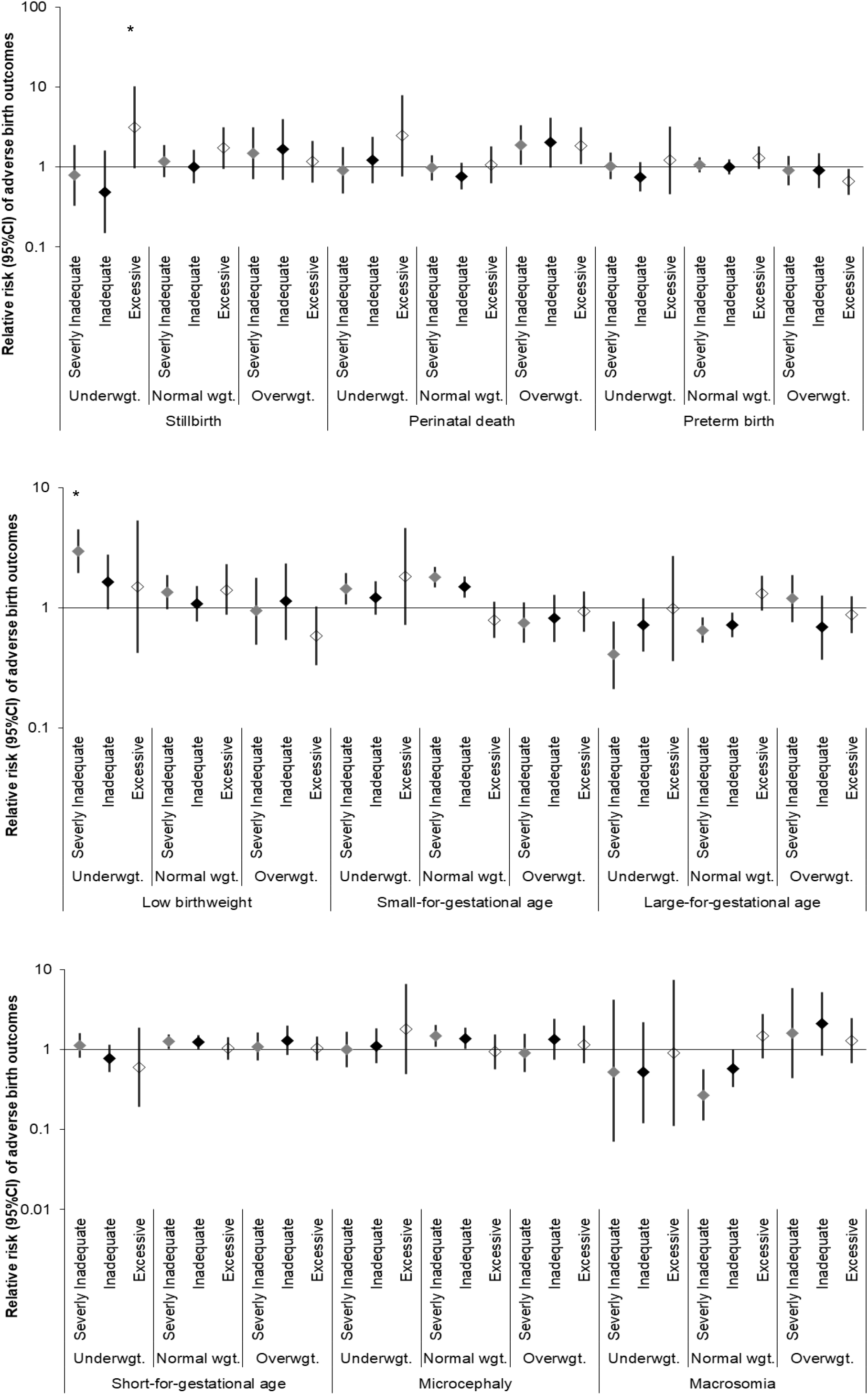
Relationship between total percent adequacy of gestational weight gain, based on Institute of Medicine recommendations, and risk of adverse neonatal outcomes, disaggregated by maternal pre-pregnancy body mass index (BMI) of underweight (<18.5 kg/m3), normal weight (18.5 – 24.9 kg/m2), and overweight or obese (≥ 25 kg/m2). An asterisk marks statistically significant (P<0.05) difference in the relationship between adequacy of gestational weight gain and neonatal outcome among women who were underweight or overweight (compared to normal weight) at the start of pregnancy.

In secondary analyses, we also examined associations between GWG z-scores and neonatal outcomes using IG-GWG standard among normal weight women (Supplementary Table 5). Relationships between total GWG z-scores and neonatal outcomes were similar to GWG adequacy ratio, with a few exceptions. For example, total GWG z-score ≥1SD was positively associated with the risk of stillbirth (RR 2.51, 95%CI: 1.26, 4.99) and GWG z-score <-1 SD in the second trimester, but not total, was positively associated with the risk of preterm birth (GWG z-score -2 SD to < -1 SD: RR 1.58, 95%CI: 1.22, 2.06; GWG z-score <-2 SD: RR 1.51, 95% CI: 1.01, 2.24) (Supplementary Table 5).

## DISCUSSION

We used secondary data from a large prospective cohort of pregnant women in Dar es Salaam, Tanzania, with multiple weight measures throughout pregnancy, to examine the relationships between GWG adequacy and a wide range of neonatal outcomes. Only a third of women in this study achieved adequate GWG based on IOM recommendations; most women had inadequate or severely inadequate GWG. Overall, inadequate GWG was associated with an increased risk of suboptimal newborn anthropometry, including LBW, SGA, stunting and microcephaly, whereas excessive GWG was positively associated the risk of LGA and macrosomia. Relationships were generally similar with GWG adequacy in the second trimester, except for a higher risk of preterm birth among women with severely inadequate GWG in the second trimester. We also observed a higher risk of stillbirths associated with excessive GWG among underweight women, compared to normal weight women.

These findings confirm the importance of GWG adequacy for neonatal health. The association of inadequate GWG with lower birthweight and a higher risk of fetal growth restriction, as measured by SGA, and of excessive GWG with LGA and macrosomia, observed in this study are consistent with previous evidence from high-, middle- and low-income settings.[2,12,20–25] Measures of size at birth have important consequences for long-term child health. Poor fetal growth is associated with an increased risk of mortality, morbidity, and adverse neurodevelopmental consequences.[4,7,26] On the other hand, excessive fetal growth is associated with poor maternal perinatal outcomes (e.g., cesarean section) and childhood obesity.[1,27] The associations of GWG with birth length and head circumference, however, have rarely been evaluated.[28–30] Inadequate GWG in this study was positively associated with the risk of stunting and microcephaly at birth. A longitudinal study of 670 pregnant women in The Gambia similarly found that greater GWG at any level was positively associated with head circumference, but only greater GWG above a threshold of 0.5 SD of conditional weight gain (i.e. greater than expected change in weight in a 3 month interval) was associated with higher birth weight and length; suggesting that better nutrition (as measured by higher GWG) may be prioritized for brain growth over other anthropometric parameters.[21] Secondary data from analysis of a multi-country prenatal nutrition supplementation trial conducted in the Democratic Republic of the Congo, Guatemala, India and Pakistan also found that higher GWG in the first trimester and higher GWG velocity overall were positively associated with birth length and weight.[28]

We also found that inadequate GWG in the second trimester was positively associated with the risk of preterm birth, an association that was also observed among normal weight women when using GWG z-scores based on the INTERGROWTH-21^st^ GWG standards. Higher GWG in the first and second trimesters, but not higher total GWG, has been previously shown to reduce the risk of spontaneous preterm birth among multigravida women.[31] Other studies have also found an increased risk of preterm birth associated with total GWG below the IOM guidelines.[2,24] The mechanisms behind the relationships between timing and degree of inadequate GWG and preterm birth require further investigation; however, it is hypothesized that inadequate GWG may be a marker for macro- and micronutrient deficiencies resulting in preterm birth, particularly if nutritional insults occur early in pregnancy which could affect plasma volume expansion or lead to inadequate maternal tissue development to support the fetus until term.[32]

In line with findings from previous studies, we found that maternal pre-pregnancy BMI modified relationships between GWG and neonatal outcomes.[2,24,30,31] Excessive GWG was positively associated with the risk of stillbirth among underweight women and among normal weight women using GWG z-scores. In addition, maternal overweight and obesity appeared to be associated with an increased risk of perinatal mortality irrespective of GWG adequacy. To the best of our knowledge, these findings are novel that have not been previously reported in the context of sub-Saharan Africa. Studies using birth cohort data in Denmark, and birth and death registration records in the United States, have also shown a higher risk of stillbirth and neonatal mortality associated with maternal overweight and obesity.[33,34] Placental dysfunction, inflammation, metabolic abnormalities, and intrapartum events are the most commonly cited mechanisms contributing to the risk of stillbirth with excessive weight.[35] However, further research is needed to understand these mechanisms and identify the contributing factors in low-income settings.

The findings of this study should be interpreted in the context of its strength and limitations. We examined the relationships between GWG adequacy and a wide range of neonatal mortality and anthropometric outcomes using one of the largest pregnancy cohorts in sub-Saharan Africa. We used GWG adequacy ratio as the primary metric as it confers the major advantage of being independent of gestational duration. Other measures of GWG, such as total absolute weight gain (in kg) and rate of weight gain (kg/week), are susceptible to confounding by gestational duration and therefore were not used.[36,37] We also used the IG-GWG among normal weight women to re-examine the evidence on GWG and adverse neonatal outcomes based on a geographically diverse reference population. However, we did not have maternal weight in the preconception period and only a few participants had weight measured in the first trimester of pregnancy; as such, we used imputed weights to assess maternal pre-pregnancy BMI and GWG. Nonetheless, we previously validated the imputation method and used the most flexible approach with the smallest imputation error to ensure the validity of our findings.[14] In addition, gestational age was based on date of the last menstrual period due to the lack of ultrasound-based assessment, the gold-standard method, in low-income settings; therefore, we cannot rule out the risk of measurement error. Finally, we used secondary data from a prenatal vitamin supplementation trial conducted in HIV-negative pregnant women, which may influence the generalizability of findings at the population level.

The World Health Organization Antenatal Guidelines recommend routinely monitoring weight during pregnancy for optimal maternal and newborn outcomes.[38] The findings from this study reaffirm the importance of optimal maternal GWG for several adverse neonatal outcomes. Most women in this study gained severely inadequate or inadequate GWG which was associated with poor newborn anthropometry, indicative of fetal growth restriction. In contrast, excessive GWG was positively associated with large size at birth and the risk of stillbirth among underweight and normal weight women. In low-income settings, multiple environmental, socio-demographic, and nutritional factors may be contributing to inadequate GWG as well as adverse neonatal outcomes [1,39]. Therefore, further research is needed to identify context-specific factors for suboptimal GWG and to identify interventions to mitigate inadequate or excessive GWG among women in Tanzania. The findings of this study reveal that greater efforts are needed to support women to achieve optimal weight pre-conceptionally and during routine antenatal care to minimize the risk of adverse neonatal outcomes.

## Data Availability

The data that support the findings of this study are not publicly available due to ethical reasons.

## Acknowledgements

We thank the mothers and children, the field teams, including nurses, midwives, supervisors, the laboratory staff, the administrative staff, and all other members of the Harvard-Tanzania collaboration for making this study possible.

## Statement of Ethics

Trial protocol was approved by the Institutional Reviews Boards at the Harvard T.H Chan School of Public Health and Muhimbili University of Health and Allied Sciences (MUHAS) in Tanzania. Only de-identified secondary data was used in this study and therefore exempt from full review by the Institutional Review Board of the Harvard T.H. Chan School of Public Health.

## Author Contributions

NP and WF conceptualized and designed the study. NP carried out the analysis and data interpretation, drafted the initial manuscript, and reviewed and revised the manuscript. DQ, AM, MW, and EL contributed to the study design, analysis, and interpretation of the data, and critically reviewed the manuscript for important intellectual content. AP and WU were investigators on the parent trial, contributed to the design and acquisition of the data, and critically reviewed the manuscript for important intellectual content. WWF was the principal investigator on the parent trial, contributed to the analysis and interpretation of the data, and critically reviewed the manuscript for important intellectual content. All authors approved the submitted manuscript.

## Data Availability Statement

The data that support the findings of this study are not publicly available due to ethical reasons.

