## Supplementary Online Material for "Associations between gestational weight gain adequacy and neonatal outcomes in Tanzania"

**Supplemental Table 1:** Associations between percent adequacy of total gestational weight gain and perinatal outcomes, modified by maternal pre-pregnancy BMI[1].

| Characteristic | n | Severely Inadequate (<70%) | | Inadequate (70% to <90%) | | Adequate (90% to 125%) | Excessive (>125%) | |
|---|---|---|---|---|---|---|---|---|
| | | RR (95% CI) | *P int*[2] | RR (95% CI) | *P int.*[2] | RR (95% CI) | RR (95% CI) | *P int.*[2] |
| **Stillbirth** | 6,579 | | | | | | | |
| Underweight (BMI <18.5 kg/m$^2$) | | 0.79 (0.33, 1.88) | 0.52 | 0.48 (0.15, 1.59) | 0.95 | 1.0 | 3.13 (0.96, 10.20) | **0.04** |
| Normal weight (18.5 – 24.9 kg/m$^2$) | | 1.17 (0.74, 1.87) | Ref. | 1.01 (0.63, 1.63) | Ref. | 1.0 | 1.73 (0.95, 3.15) | Ref. |
| Overweight/Obese (≥ 25 kg/m$^2$) | | 1.49 (0.71, 3.13) | 0.81 | 1.66 (0.69, 4.00) | 0.99 | 1.0 | 1.17 (0.64, 2.12) | 0.40 |
| **Perinatal death** | 6,376 | | | | | | | |
| Underweight (BMI <18.5 kg/m$^2$) | | 0.91 (0.47, 1.78) | 0.64 | 1.23 (0.63, 2.40) | 0.31 | 1.0 | 2.45 (0.76, 7.93) | 0.09 |
| Normal weight (18.5 – 24.9 kg/m$^2$) | | 0.98 (0.68, 1.40) | Ref. | 0.76 (0.52, 1.12) | Ref. | 1.0 | 1.06 (0.63, 1.80) | Ref. |
| Overweight/Obese (≥ 25 kg/m$^2$) | | 1.87 (1.06, 3.30) | 0.55 | 2.02 (0.99, 4.09) | 0.49 | 1.0 | 1.84 (1.09, 3.12) | 0.56 |
| **Preterm-born** | 6,370 | | | | | | | |
| Underweight (BMI <18.5 kg/m$^2$) | | 1.03 (0.71, 1.51) | 0.53 | 0.75 (0.49, 1.16) | 0.72 | 1.0 | 1.21 (0.46, 3.18) | 0.52 |
| Normal weight (18.5 – 24.9 kg/m$^2$) | | 1.06 (0.85, 1.32) | Ref. | 1.00 (0.80, 1.24) | Ref. | 1.0 | 1.30 (0.94, 1.81) | Ref. |
| Overweight/Obese (≥ 25 kg/m$^2$) | | 0.90 (0.59, 1.37) | 0.40 | 0.90 (0.54, 1.49) | 0.45 | 1.0 | 0.66 (0.45, 0.94) | 0.78 |
| **Low birthweight** | 6,205 | | | | | | | |
| Underweight (BMI <18.5 kg/m$^2$) | | 2.95 (1.95, 4.48) | **0.03** | 1.65 (0.97, 2.78) | 0.46 | 1.0 | 1.49 (0.42, 5.33) | 0.76 |
| Normal weight (18.5 – 24.9 kg/m$^2$) | | 1.34 (0.97, 1.87) | Ref. | 1.08 (0.77, 1.52) | Ref. | 1.0 | 1.41 (0.87, 2.30) | Ref. |
| Overweight/Obese (≥ 25 kg/m$^2$) | | 0.94 (0.49, 1.78) | 0.45 | 1.13 (0.54, 2.35) | 0.28 | 1.0 | 0.58 (0.33, 1.02) | 0.77 |
| **Small-for-gestational age** | 6,188 | | | | | | | |
| Underweight (BMI <18.5 kg/m$^2$) | | 1.44 (1.07, 1.95) | 0.23 | 1.21 (0.87, 1.67) | 0.59 | 1.0 | 1.83 (0.72, 4.64) | 0.28 |
| Normal weight (18.5 – 24.9 kg/m$^2$) | | 1.79 (1.47, 2.18) | Ref. | 1.49 (1.22, 1.81) | Ref. | 1.0 | 0.79 (0.56, 1.12) | Ref. |
| Overweight/Obese (≥ 25 kg/m$^2$) | | 0.75 (0.51, 1.11) | 0.40 | 0.82 (0.52, 1.28) | 0.63 | 1.0 | 0.93 (0.63, 1.36) | 0.97 |
| **Large-for-gestational age** | 6,188 | | | | | | | |
| Underweight (BMI <18.5 kg/m$^2$) | | 0.41 (0.21, 0.77) | 0.82 | 0.72 (0.43, 1.19) | 0.27 | 1.0 | 0.99 (0.36, 2.71) | 0.20 |
| Normal weight (18.5 – 24.9 kg/m$^2$) | | 0.65 (0.51, 0.83) | Ref. | 0.72 (0.57, 0.91) | Ref. | 1.0 | 1.32 (0.95, 1.84) | Ref. |
| Overweight/Obese (≥ 25 kg/m$^2$) | | 1.20 (0.76, 1.88) | 0.08 | 0.69 (0.37, 1.27) | 0.91 | 1.0 | 0.87 (0.61, 1.25) | 0.44 |
| **Stunting at birth** | 4,590 | | | | | | | |
| Underweight (BMI <18.5 kg/m$^2$) | | 1.12 (0.79, 1.60) | 0.91 | 0.77 (0.52, 1.14) | 0.16 | 1.0 | 0.60 (0.19, 1.89) | 0.29 |
| Normal weight (18.5 – 24.9 kg/m$^2$) | | 1.26 (1.02, 1.55) | Ref. | 1.23 (1.00, 1.51) | Ref. | 1.0 | 1.03 (0.75, 1.42) | Ref. |
| Overweight/Obese (≥ 25 kg/m$^2$) | | 1.09 (0.73, 1.62) | 0.96 | 1.30 (0.85, 2.00) | 0.46 | 1.0 | 1.03 (0.73, 1.44) | 0.85 |
| **Microcephaly** | 5,435 | | | | | | | |
| Underweight (BMI <18.5 kg/m$^2$) | | 1.00 (0.60, 1.66) | 0.38 | 1.10 (0.67, 1.82) | 0.54 | 1.0 | 1.81 (0.49, 6.61) | 0.74 |
| Normal weight (18.5 – 24.9 kg/m$^2$) | | 1.48 (1.09, 2.01) | Ref. | 1.37 (1.01, 1.86) | Ref. | 1.0 | 0.94 (0.56, 1.55) | Ref. |
| Overweight/Obese (≥ 25 kg/m$^2$) | | 0.90 (0.52, 1.58) | 0.32 | 1.34 (0.75, 2.41) | 0.93 | 1.0 | 1.15 (0.67, 1.97) | 0.73 |
| **Macrosomia** | 6,205 | | | | | | | |
| Underweight (BMI <18.5 kg/m$^2$) | | 0.52 (0.07, 4.20) | 0.86 | 0.52 (0.12, 2.19) | 0.80 | 1.0 | 0.90 (0.11, 7.38) | 0.80 |
| Normal weight (18.5 – 24.9 kg/m$^2$) | | 0.27 (0.13, 0.56) | Ref. | 0.58 (0.34, 1.00) | Ref. | 1.0 | 1.47 (0.78, 2.77) | Ref. |
| Overweight/Obese (≥ 25 kg/m$^2$) | | 1.61 (0.44, 5.87) | 0.60 | 2.11 (0.84, 5.25) | 0.24 | 1.0 | 1.29 (0.68, 2.45) | 0.71 |

[1] Multivariable models adjusted for maternal age, education, marital status, wealth quintile, smoking status, alcohol consumption, maternal pre-pregnancy BMI, and supplementation group assignment.
[2] P-value for interaction based on Wald test evaluating the relationship between gestational weight gain adequacy ratio and perinatal outcome among normal weight women is the reference.

**Supplemental Table 2:** Associations between percent adequacy of early gestational weight gain (2nd trimester) and perinatal outcomes, modified by maternal pre-pregnancy BMI[1].

| Characteristic | n | Severely Inadequate (<70%) | | Inadequate (70% to <90%) | | Adequate (90% to 125%) | Excessive (>125%) | |
|---|---|---|---|---|---|---|---|---|
| | | RR (95% CI) | *P int*[2] | RR (95% CI) | *P int.*[2] | RR (95% CI) | RR (95% CI) | *P int.*[2] |
| **Stillbirth** | 7,054 | | | | | | | |
| Underweight (BMI <18.5 kg/m$^2$) | | 0.97 (0.33, 2.83) | 0.49 | 0.71 (0.33, 1.51) | 0.72 | 1.0 | 1.90 (0.66, 5.44) | 0.12 |
| Normal weight (18.5 – 24.9 kg/m$^2$) | | 0.94 (0.58, 1.53) | Ref. | 1.07 (0.72, 1.59) | Ref. | 1.0 | 1.14 (0.64, 2.03) | Ref. |
| Overweight/Obese (≥ 25 kg/m$^2$) | | 2.06 (1.00, 4.23) | 0.63 | 0.70 (0.17, 2.91) | 0.31 | 1.0 | 1.55 (0.88, 2.73) | 0.93 |
| **Perinatal death** | 6,750 | | | | | | | |
| Underweight (BMI <18.5 kg/m$^2$) | | 1.36 (0.64, 2.90) | 0.65 | 0.84 (0.47, 1.50) | 0.58 | 1.0 | 1.26 (0.49, 3.23) | 0.79 |
| Normal weight (18.5 – 24.9 kg/m$^2$) | | 0.94 (0.64, 1.37) | Ref. | 0.92 (0.67, 1.26) | Ref. | 1.0 | 1.13 (0.72, 1.78) | Ref. |
| Overweight/Obese (≥ 25 kg/m$^2$) | | 2.26 (1.30, 3.93) | 0.38 | 1.80 (0.83, 3.89) | 0.80 | 1.0 | 1.52 (0.97, 2.37) | 0.90 |
| **Preterm-born** | 6,807 | | | | | | | |
| Underweight (BMI <18.5 kg/m$^2$) | | 1.11 (0.71, 1.72) | 0.17 | 0.77 (0.53, 1.11) | 0.75 | 1.0 | 1.12 (0.59, 2.12) | 0.24 |
| Normal weight (18.5 – 24.9 kg/m$^2$) | | 1.55 (1.25, 1.91) | Ref. | 0.86 (0.71, 1.05) | Ref. | 1.0 | 1.17 (0.89, 1.56) | Ref. |
| Overweight/Obese (≥ 25 kg/m$^2$) | | 0.80 (0.54, 1.19) | 0.47 | **2.19 (1.32, 3.62)** | **0.02** | 1.0 | 0.62 (0.45, 0.84) | 0.07 |
| **Low birthweight** | 6,579 | | | | | | | |
| Underweight (BMI <18.5 kg/m$^2$) | | 3.26 (1.94, 5.45) | 0.02 | 1.61 (1.05, 2.45) | 0.54 | 1.0 | 1.63 (0.70, 3.81) | 0.64 |
| Normal weight (18.5 – 24.9 kg/m$^2$) | | 1.43 (1.03, 2.00) | Ref. | 1.16 (0.87, 1.56) | Ref. | 1.0 | 1.26 (0.81, 1.95) | Ref. |
| Overweight/Obese (≥ 25 kg/m$^2$) | | 1.05 (0.58, 1.91) | 0.94 | 1.14 (0.48, 2.68) | 0.94 | 1.0 | 0.68 (0.42, 1.09) | 0.23 |
| **Small-for-gestational age** | 6,559 | | | | | | | |
| Underweight (BMI <18.5 kg/m$^2$) | | 1.31 (0.86, 2.00) | 1.0 | 1.34 (1.03, 1.76) | 0.92 | 1.0 | 1.34 (0.72, 2.50) | 0.96 |
| Normal weight (18.5 – 24.9 kg/m$^2$) | | 1.28 (1.04, 1.58) | Ref. | 1.38 (1.15, 1.64) | Ref. | 1.0 | 0.82 (0.61, 1.10) | Ref. |
| Overweight/Obese (≥ 25 kg/m$^2$) | | 0.83 (0.55, 1.25) | 0.96 | 0.57 (0.30, 1.09) | 0.32 | 1.0 | 0.91 (0.66, 1.26) | 0.77 |
| **Large-for-gestational age** | 6,559 | | | | | | | |
| Underweight (BMI <18.5 kg/m$^2$) | | 0.58 (0.32, 1.07) | 0.48 | 0.57 (0.35, 0.93) | 0.46 | 1.0 | 0.52 (0.22, 1.22) | 0.70 |
| Normal weight (18.5 – 24.9 kg/m$^2$) | | 1.23 (0.98, 1.56) | Ref. | 0.74 (0.59, 0.92) | Ref. | 1.0 | 1.28 (0.95, 1.72) | Ref. |
| Overweight/Obese (≥ 25 kg/m$^2$) | | 0.83 (0.54, 1.27) | 0.53 | 1.79 (0.98, 3.26) | 0.13 | 1.0 | 0.85 (0.63, 1.16) | 0.55 |
| **Stunting at birth** | 4,741 | | | | | | | |
| Underweight (BMI <18.5 kg/m$^2$) | | 0.73 (0.42, 1.24) | 0.13 | 1.12 (0.82, 1.52) | 0.70 | 1.0 | 0.67 (0.34, 1.34) | 0.14 |
| Normal weight (18.5 – 24.9 kg/m$^2$) | | 1.14 (0.91, 1.44) | Ref. | 1.09 (0.90, 1.33) | Ref. | 1.0 | 1.17 (0.88, 1.55) | Ref. |
| Overweight/Obese (≥ 25 kg/m$^2$) | | 0.69 (0.44, 1.08) | 0.04 | 0.88 (0.47, 1.65) | 0.33 | 1.0 | 0.97 (0.72, 1.30) | 0.27 |
| **Microcephaly** | 5,615 | | | | | | | |
| Underweight (BMI <18.5 kg/m$^2$) | | 0.79 (0.37, 1.68) | 0.43 | 1.33 (0.86, 2.04) | 0.74 | 1.0 | 1.35 (0.56, 3.23) | 0.79 |
| Normal weight (18.5 – 24.9 kg/m$^2$) | | 1.09 (0.78, 1.51) | Ref. | 1.02 (0.77, 1.35) | Ref. | 1.0 | 0.98 (0.65, 1.49) | Ref. |
| Overweight/Obese (≥ 25 kg/m$^2$) | | 0.93 (0.50, 1.75) | 0.66 | 0.50 (0.15, 1.61) | 0.22 | 1.0 | 1.06 (0.68, 1.64) | 0.86 |
| **Macrosomia** | 6,579 | | | | | | | |
| Underweight (BMI <18.5 kg/m$^2$) | | 0.95 (0.11, 7.99) | 0.82 | 0.45 (0.11, 1.91) | 0.62 | 1.0 | 0.29 (0.04, 2.20) | 0.44 |
| Normal weight (18.5 – 24.9 kg/m$^2$) | | 0.35 (0.16, 0.79) | Ref. | 0.59 (0.34, 1.00) | Ref. | 1.0 | 2.10 (1.22, 3.62) | Ref. |
| Overweight/Obese (≥ 25 kg/m$^2$) | | 1.66 (0.43, 6.36) | 0.39 | 2.29 (0.65, 8.06) | 0.20 | 1.0 | 0.99 (0.58, 1.67) | 0.72 |

[1] Multivariable models adjusted for maternal age, education, marital status, wealth quintile, smoking status, alcohol consumption, maternal pre-pregnancy BMI, and supplementation group assignment.
[2] P-value for interaction based on Wald test evaluating the relationship between gestational weight gain adequacy ratio and perinatal outcome among normal weight women is the reference.

**Supplemental Table 3:** Associations between percent adequacy of total gestational weight gain and perinatal outcomes, modified by maternal age[1].

| Characteristic | n | Severely Inadequate (<70%) | | Inadequate (70% to <90%) | | Adequate (90% to 125%) | Excessive (>125%) | |
|---|---|---|---|---|---|---|---|---|
| | | RR (95% CI) | *P int*[2] | RR (95% CI) | *P int.*[2] | RR (95% CI) | RR (95% CI) | *P int.*[2] |
| **Stillbirth** | 6,579 | | | | | | | |
| <20 years | | 1.24 (0.55, 2.82) | 0.76 | 0.78 (0.32, 1.86) | 0.65 | 1.0 | 1.85 (0.84, 4.04) | 0.32 |
| 20 – 29 years | | 1.03 (0.51, 2.06) | Ref. | 1.38 (0.72, 2.65) | Ref. | 1.0 | 1.70 (0.84, 3.48) | Ref. |
| >30 years | | 2.12 (1.09, 4.14) | 0.61 | 0.82 (0.40, 1.70) | 0.12 | 1.0 | 1.23 (0.65, 2.32) | 0.46 |
| **Perinatal death** | 6,376 | | | | | | | |
| <20 years | | 1.06 (0.59, 1.94) | 0.65 | 0.74 (0.37, 1.47) | 0.71 | 1.0 | 1.57 (0.77, 3.17) | 0.22 |
| 20 – 29 years | | 0.90 (0.56, 1.45) | Ref. | 0.99 (0.62, 1.58) | Ref. | 1.0 | 0.92 (0.54, 1.56) | Ref. |
| >30 years | | 1.42 (0.85, 2.38) | 0.44 | 0.77 (0.44, 1.37) | 0.32 | 1.0 | 1.64 (0.97, 2.77) | 0.24 |
| **Preterm-born** | 6,370 | | | | | | | |
| <20 years | | 0.93 (0.64, 1.37) | 0.049 | 1.19 (0.79, 1.80) | 0.30 | 1.0 | 1.66 (0.99, 2.81) | 0.89 |
| 20 – 29 years | | 1.25 (0.93, 1.69) | Ref. | 1.01 (0.75, 1.38) | Ref. | 1.0 | 1.27 (0.88, 1.84) | Ref. |
| >30 years | | 0.75 (0.54, 1.04) | 0.9 | 0.86 (0.62, 1.21) | 0.6 | 1.0 | 0.71 (0.48, 1.06) | 0.74 |
| **Low birthweight** | 6,205 | | | | | | | |
| <20 years | | 1.00 (0.62, 1.62) | 1.0 | 1.20 (0.69, 2.09) | 0.66 | 1.0 | 1.18 (0.50, 2.76) | 0.77 |
| 20 – 29 years | | 1.84 (1.19, 2.85) | Ref. | 1.31 (0.83, 2.09) | Ref. | 1.0 | 1.22 (0.66, 2.22) | Ref. |
| >30 years | | 0.88 (0.56, 1.38) | 0.36 | 0.78 (0.47, 1.29) | 0.23 | 1.0 | 1.26 (0.69, 2.30) | 0.85 |
| **Small-for-gestational age** | 6,188 | | | | | | | |
| <20 years | | 1.15 (0.85, 1.54) | 0.15 | 1.24 (0.89, 1.73) | 0.09 | 1.0 | 0.87 (0.50, 1.53) | 0.83 |
| 20 – 29 years | | 1.71 (1.32, 2.21) | Ref. | 1.44 (1.10, 1.87) | Ref. | 1.0 | 0.90 (0.64, 1.28) | Ref. |
| >30 years | | 1.04 (0.79, 1.37) | 0.97 | 0.93 (0.70, 1.24) | 0.54 | 1.0 | 0.87 (0.59, 1.29) | 0.43 |
| **Large-for-gestational age** | 6,188 | | | | | | | |
| <20 years | | 1.06 (0.66, 1.70) | 0.30 | 1.14 (0.67, 1.95) | 0.45 | 1.0 | 1.61 (0.92, 2.81) | 0.80 |
| 20 – 29 years | | 0.85 (0.61, 1.19) | Ref. | 0.72 (0.51, 1.02) | Ref. | 1.0 | 1.43 (0.99, 2.08) | Ref. |
| >30 years | | 0.60 (0.41, 0.88) | 0.14 | 1.03 (0.71, 1.49) | 0.45 | 1.0 | 0.85 (0.59, 1.23) | 0.95 |
| **Stunting at birth** | 4,590 | | | | | | | |
| <20 years | | 1.08 (0.75, 1.56) | 0.13 | 0.87 (0.58, 1.28) | 0.53 | 1.0 | 0.61 (0.35, 1.08) | 0.62 |
| 20 – 29 years | | 0.95 (0.72, 1.26) | Ref. | 1.10 (0.83, 1.46) | Ref. | 1.0 | 0.99 (0.71, 1.38) | Ref. |
| >30 years | | **1.39 (1.03, 1.89)** | **0.01** | 0.94 (0.69, 1.28) | 0.57 | 1.0 | 0.83 (0.59, 1.18) | 0.96 |
| **Microcephaly** | 5,435 | | | | | | | |
| <20 years | | 1.47 (0.92, 2.35) | 0.07 | 0.89 (0.52, 1.54) | 0.63 | 1.0 | 1.20 (0.58, 2.51) | 0.30 |
| 20 – 29 years | | 1.04 (0.68, 1.58) | Ref. | 1.19 (0.79, 1.79) | Ref. | 1.0 | 0.89 (0.53, 1.49) | Ref. |
| >30 years | | 1.41 (0.91, 2.18) | 0.29 | 1.23 (0.81, 1.85) | 0.55 | 1.0 | 0.92 (0.53, 1.59) | 0.72 |
| **Macrosomia** | 6,205 | | | | | | | |
| <20 years | | 0.80 (0.16, 3.98) | 0.46 | 1.77 (0.52, 6.01) | 0.91 | 1.0 | 1.74 (0.58, 5.20) | 0.92 |
| 20 – 29 years | | 0.69 (0.27, 1.77) | Ref. | 0.76 (0.31, 1.87) | Ref. | 1.0 | 1.81 (0.79, 4.15) | Ref. |
| >30 years | | **0.45 (0.13, 1.54)** | **0.03** | 1.69 (0.70, 4.08) | 0.64 | 1.0 | 1.67 (0.85, 3.28) | 0.56 |

[1]Multivariable models adjusted for maternal age, education, marital status, wealth quintile, smoking status, alcohol consumption, maternal pre-pregnancy BMI, and supplementation group assignment.
[2] P-value for interaction based on Wald test evaluating the relationship between gestational weight gain adequacy ratio and perinatal outcome among normal weight women is the reference.

**Supplemental Table 4:** Associations between percent adequacy of early gestational weight gain (2nd trimester) and perinatal outcomes, modified by maternal age[1].

| Characteristic | n | Severely Inadequate (<70%) | | Inadequate (70% to <90%) | | Adequate (90% to 125%) | Excessive (>125%) | |
|---|---|---|---|---|---|---|---|---|
| | | RR (95% CI) | *P int*[2] | RR (95% CI) | *P int.*[2] | RR (95% CI) | RR (95% CI) | *P int.*[2] |
| **Stillbirth** | 7,054 | | | | | | | |
| <20 years | | 0.69 (0.28, 1.66) | 0.96 | 1.88 (0.96, 3.67) | 0.06 | 1.0 | 1.23 (0.58, 2.64) | 0.31 |
| 20 – 29 years | | 1.06 (0.59, 1.92) | Ref. | 0.67 (0.37, 1.20) | Ref. | 1.0 | 0.97 (0.52, 1.81) | Ref. |
| >30 years | | 0.96 (0.47, 1.95) | 0.94 | 1.71 (0.94, 3.10) | 0.18 | 1.0 | 1.38 (0.79, 2.39) | 0.38 |
| **Perinatal death** | 6,750 | | | | | | | |
| <20 years | | 1.07 (0.57, 2.01) | 0.34 | 1.39 (0.81, 2.39) | 0.09 | 1.0 | 0.94 (0.50, 1.77) | 0.51 |
| 20 – 29 years | | 0.86 (0.54, 1.38) | Ref. | 0.68 (0.44, 1.05) | Ref. | 1.0 | 0.94 (0.59, 1.50) | Ref. |
| >30 years | | 1.25 (0.72, 2.17) | 0.41 | 1.36 (0.84, 2.19) | 0.24 | 1.0 | 1.11 (0.73, 1.71) | 0.56 |
| **Preterm-born** | 6,807 | | | | | | | |
| <20 years | | 1.02 (0.70, 1.48) | 0.22 | 1.23 (0.87, 1.75) | 0.63 | 1.0 | 1.26 (0.81, 1.97) | 0.73 |
| 20 – 29 years | | 1.56 (1.17, 2.07) | Ref. | 0.88 (0.67, 1.15) | Ref. | 1.0 | 1.07 (0.77, 1.47) | Ref. |
| >30 years | | 0.89 (0.64, 1.22) | 0.33 | 0.80 (0.59, 1.08) | 0.61 | 1.0 | 0.64 (0.46, 0.90) | 0.60 |
| **Low birthweight** | 6,579 | | | | | | | |
| <20 years | | 1.05 (0.63, 1.75) | 0.94 | 1.19 (0.75, 1.89) | 0.78 | 1.0 | 0.94 (0.45, 1.97) | 0.75 |
| 20 – 29 years | | 1.60 (1.06, 2.43) | Ref. | 1.08 (0.73, 1.59) | Ref. | 1.0 | 0.93 (0.56, 1.55) | Ref. |
| >30 years | | 0.95 (0.59, 1.53) | 0.68 | 0.98 (0.64, 1.50) | 0.59 | 1.0 | 1.26 (0.77, 2.08) | 0.21 |
| **Small-for-gestational age** | 6,559 | | | | | | | |
| <20 years | | 1.09 (0.77, 1.56) | 0.41 | 1.17 (0.88, 1.56) | 0.22 | 1.0 | 1.09 (0.71, 1.69) | 0.47 |
| 20 – 29 years | | 1.24 (0.94, 1.63) | Ref. | 1.22 (0.96, 1.56) | Ref. | 1.0 | 0.86 (0.63, 1.16) | Ref. |
| >30 years | | 0.92 (0.66, 1.29) | 0.9 | 1.10 (0.85, 1.41) | 0.41 | 1.0 | 0.89 (0.64, 1.23) | 0.77 |
| **Large-for-gestational age** | 6,559 | | | | | | | |
| <20 years | | 0.94 (0.59, 1.49) | 0.75 | 1.71 (1.13, 2.58) | 0.09 | 1.0 | 1.54 (0.95, 2.49) | 0.22 |
| 20 – 29 years | | 1.27 (0.92, 1.75) | Ref. | 0.68 (0.50, 0.94) | Ref. | 1.0 | 1.10 (0.78, 1.55) | Ref. |
| >30 years | | 0.75 (0.52, 1.09) | 0.91 | 0.84 (0.59, 1.20) | 0.7 | 1.0 | 0.81 (0.58, 1.12) | 0.84 |
| **Stunting at birth** | 4,741 | | | | | | | |
| <20 years | | 1.10 (0.72, 1.68) | 0.27 | 0.85 (0.60, 1.20) | 0.82 | 1.0 | 0.63 (0.39, 1.03) | 0.46 |
| 20 – 29 years | | 0.87 (0.64, 1.19) | Ref. | 0.92 (0.71, 1.20) | Ref. | 1.0 | 1.05 (0.77, 1.44) | Ref. |
| >30 years | | 0.97 (0.67, 1.40) | 0.52 | 1.13 (0.86, 1.50) | 0.12 | 1.0 | 0.94 (0.70, 1.26) | 0.58 |
| **Microcephaly** | 5,615 | | | | | | | |
| <20 years | | 1.15 (0.66, 2.03) | 0.17 | 1.24 (0.77, 1.99) | 0.09 | 1.0 | 1.22 (0.65, 2.32) | 0.16 |
| 20 – 29 years | | 0.99 (0.64, 1.53) | Ref. | 0.87 (0.59, 1.29) | Ref. | 1.0 | 0.85 (0.53, 1.34) | Ref. |
| >30 years | | 0.90 (0.52, 1.56) | 0.53 | 1.24 (0.82, 1.86) | 0.7 | 1.0 | 1.33 (0.84, 2.10) | 0.54 |
| **Macrosomia** | 6,579 | | | | | | | |
| <20 years | | 1.06 (0.20, 5.52) | 0.73 | 1.55 (0.52, 4.69) | 0.37 | 1.0 | 2.19 (0.87, 5.51) | 0.17 |
| 20 – 29 years | | 0.63 (0.22, 1.80) | Ref. | 0.77 (0.33, 1.78) | Ref. | 1.0 | 1.58 (0.70, 3.58) | Ref. |
| >30 years | | 0.73 (0.19, 2.82) | 0.22 | 1.07 (0.44, 2.58) | 0.33 | 1.0 | 2.18 (1.20, 3.99) | 0.71 |

[1]Multivariable models adjusted for maternal age, education, marital status, wealth quintile, smoking status, alcohol consumption, maternal pre-pregnancy BMI, and supplementation group assignment.
[2]P-value for interaction based on Wald test evaluating the relationship between gestational weight gain adequacy ratio and perinatal outcome among normal weight women is the reference.

**Supplementary Table 5:** Associations between gestational weight gain (GWG) z-scores, using Intergrowth-21st standards, and perinatal outcomes, among normal weight women[1].

| Characteristic | Total n | No. of cases (%) | GWGz <-2 SD RR (95% CI) | GWGz -2 SD to <-1 SD RR (95% CI) | GWGz -1 SD to <1 SD Reference | GWGz ≥1 SD RR (95% CI) |
|---|---|---|---|---|---|---|
| **Total GWG** | | | | | | |
| Stillbirth | 4314 | 123 (2.85) | 1.27 (0.65, 2.48) | 0.98 (0.63, 1.54) | 1.0 | 2.51 (1.26, 4.99) |
| Perinatal death | 4165 | 195 (4.68) | 1.06 (0.60, 1.87) | 0.94 (0.66, 1.35) | 1.0 | 1.45 (0.73, 2.85) |
| Preterm-born | 4191 | 630 (15.0) | 0.82 (0.57, 1.18) | 0.80 (0.65, 0.98) | 1.0 | 1.48 (0.97, 2.25) |
| Low birthweight | 4076 | 243 (5.96) | 1.62 (1.02, 2.55) | 0.93 (0.68, 1.29) | 1.0 | 1.43 (0.73, 2.77) |
| Small-for-gestational age | 4066 | 866 (21) | 1.72 (1.31, 2.27) | 1.54 (1.30, 1.83) | 1.0 | 0.73 (0.45, 1.18) |
| Large-for-gestational age | 4066 | 512 (12.6) | 0.51 (0.33, 0.81) | 0.62 (0.49, 0.79) | 1.0 | 1.52 (0.97, 2.37) |
| Stunting at birth | 2996 | 797 (27) | 1.16 (0.84, 1.59) | 1.16 (0.96, 1.41) | 1.0 | 0.64 (0.39, 1.08) |
| Microcephaly | 3564 | 307 (8.62) | 1.46 (0.96, 2.22) | 1.39 (1.06, 1.82) | 1.0 | 0.61 (0.26, 1.41) |
| Macrosomia | 4076 | 83 (2.04) | 0.16 (0.02, 1.13) | 0.37 (0.18, 0.74) | 1.0 | 1.62 (0.66, 4.01) |
| **Early GWG (2nd trimester)** | | | | | | |
| Stillbirth | 4630 | 145 (3.13) | 1.89 (0.95, 3.76) | 0.87 (0.45, 1.67) | 1.0 | 1.08 (0.60, 1.95) |
| Perinatal death | 4411 | 231 (5.24) | 1.26 (0.65, 2.42) | 0.94 (0.56, 1.57) | 1.0 | 1.22 (0.77, 1.92) |
| Preterm-born | 4485 | 753 (16.8) | 1.51 (1.01, 2.24) | 1.58 (1.22, 2.06) | 1.0 | 1.19 (0.90, 1.58) |
| Low birthweight | 4325 | 286 (6.61) | 1.36 (0.75, 2.46) | 1.30 (0.85, 1.97) | 1.0 | 1.39 (0.92, 2.10) |
| Small-for-gestational age | 4313 | 910 (21) | 1.05 (0.70, 1.56) | 1.10 (0.85, 1.43) | 1.0 | 0.63 (0.47, 0.86) |
| Large-for-gestational age | 4313 | 586 (13.6) | 1.30 (0.83, 2.05) | 1.28 (0.94, 1.73) | 1.0 | 1.51 (1.13, 2.01) |
| Stunting at birth | 3094 | 814 (26) | 1.09 (0.72, 1.67) | 1.23 (0.92, 1.66) | 1.0 | 1.23 (0.93, 1.62) |
| Microcephaly | 3679 | 312 (8.48) | 1.68 (1.01, 2.81) | 0.95 (0.61, 1.50) | 1.0 | 0.89 (0.57, 1.38) |
| Macrosomia | 4325 | 86 (1.99) | 0.82 (0.20, 3.32) | 0.46 (0.14, 1.47) | 1.0 | 2.82 (1.65, 4.80) |

[1] Multivariable models adjusted for maternal age, education, marital status, wealth quintile, smoking status, alcohol consumption, maternal pre-pregnancy body-mass-index, parity, and supplementation group assignment.
